# The impact of vaccination on incidence and outcomes of SARS-CoV-2 infection in patients with kidney failure in Scotland

**DOI:** 10.1101/2022.01.05.22268785

**Authors:** Samira Bell, Jacqueline Campbell, Emilie Lambourg, Chrissie Watters, Martin O’Neil, Alison Almond, Katharine Buck, Edward J Carr, Laura Clark, Zoe Cousland, Mark Findlay, Nicola Joss, Wendy Metcalfe, Michaela Petrie, Elaine Spalding, Jamie P Traynor, Vinod Sanu, Peter Thomson, Shona Methven, Patrick B Mark, on behalf of the Scottish Renal Registry

## Abstract

**Background:** Patients with kidney failure requiring kidney replacement therapy (KRT) are at high risk of complications and death following SARS-CoV-2 infection with variable antibody responses to vaccination reported. We investigated the effects of COVID-19 vaccination on incidence of infection, hospitalization and death of COVID-19 infection.

**Methods:** Study design was an observational data linkage cohort study. Multiple healthcare datasets were linked to ascertain all SARS-CoV-2 testing, vaccination, hospitalization, and mortality data for all patients treated with KRT in Scotland, from the start of the pandemic over a period of 20 months. Descriptive statistics, survival analyses and vaccine effectiveness were calculated.

**Results:** As of 19th September 2021, 93% (n=5281) of the established KRT population in Scotland had received two doses of an approved SARS-CoV-2 vaccine. Over the study period, there were 814 cases of SARS-CoV-2 infection (15.1% of the KRT population). Vaccine effectiveness against infection and hospitalization was 33% (95% CI 0-52) and 38% (95% CI 0-57) respectively. 9.2% of fully vaccinated individuals died within 28 days of a SARS-CoV-2 positive PCR test (7% dialysis patients and 10% kidney transplant recipients). This compares to <0.1% of the vaccinated Scottish population being admitted to hospital or dying death due to COVID19 during that period.

**Conclusions:** These data demonstrate a primary vaccine course of two doses has limited impact on COVID-19 infection and its complications in patients treated with KRT. Adjunctive strategies to reduce risk of both COVID-19 infection and its complications in this population are urgently required.

## Introduction

Patients requiring kidney replacement therapy (KRT) with dialysis or kidney transplantation are at high risk of death following SARS-CoV-2 infection, with case fatality at 28-days reported at 16.9% in kidney transplant recipients (KTR) and 23.9% in patients requiring hemodialysis (HD) in the European Renal Association COVID-19 registry. Similarly, poor outcomes are reported globally^1-4^. Vaccination against COVID-19 reduces mortality and risk of hospitalization from COVID-19 in the general population^5^. Kidney transplant recipients (KTR) on immunosuppression have been demonstrated to have attenuated serological response to conventional two-dose COVID-19 vaccination schedules^6, 7^. Serological response to vaccination in patients treated with hemodialysis (HD) is variable with most studies suggesting that vaccination induces anti-Spike antibodies^8, 9^. Many countries recognized patients undergoing HD and KTR as ‘clinically extremely vulnerable’ and prioritized early vaccination of patients requiring KRT. In Scotland, 90% of first doses of a COVID-19 vaccine were administered by 28 February 2021 in Scotland to patients requiring KRT.

The emergence of more transmissible SARS-CoV-2 variants of concern (VOC), in particular the Delta (B.1.617.2) variant, which became the dominant strain in the UK in May 2021 has driven rapid increases in the incidence of COVID-19^10^. Differences in induction of neutralizing antibodies to the Delta VOC in patients treated with HD after vaccination with BNT162b2 (Pfizer–BioNTech) or ChAdOx1 (Oxford–AstraZeneca) vaccines have been demonstrated^11^. Vaccination with two doses of BNT162b2 in patients undergoing HD induces similar neutralizing antibody titers to those in healthy individuals. However, vaccination with two doses of ChAdOx1 in COVID-19 seronaïve patients undergoing HD induces suboptimal neutralizing antibodies to the Delta variant. Therefore, there is reduced serological response to vaccination in KTR and differential efficacy of BNT162b2 and ChAdOX1 in patients undergoing HD against the dominant Delta VOC. The clinical implications of these combined findings on COVID-19 infection, hospitalization and death are unknown. We investigated the effects of SARS-CoV-2 vaccination on incidence and clinical outcomes of COVID-19.

## Methods

### Data Sources

Study design was an observational data linkage cohort study. The following national datasets were linked using a unique patient identifier (Community Health Index (CHI)) by Public Health and Intelligence unit of Public Health, Scotland. The Scottish Renal Registry is a national registry of all patients receiving KRT for kidney failure in Scotland (hemodialysis, peritoneal dialysis and transplant). It collates data from all 9 adult renal units in Scotland and 28 satellite hemodialysis units serving a population of 5.4 million. The Scottish Renal Registry was established in 1991 with data backfilled to 1960 from European Renal Association-European Dialysis and Transplant Association (ERA-EDTA) with the first patient dialyzed for kidney failure in Scotland in 1960. It has 100% unit and patient coverage. Data held by the registry include patient demographics including historical postcodes (for calculating Scottish Index of Multiple Deprivation), full KRT history (for kidney failure), primary renal diagnosis (using ERA-EDTA codes) and linkage with National Health Service Blood and Transplant (NHS BT) for transplant status. The primary renal diagnosis groupings used in the analyses is described in the Scottish Renal Registry website^12^. The Scottish Government provides online calculators allowing use of patient postcode to generate divisions of socioeconomic deprivation, the Scottish Index of Multiple Deprivation (SIMD)^13^. Using patient postcode, deprivation quintiles were calculated with 1 corresponding to most deprived and 5 to least deprived. Data on SARS-CoV-2 testing were obtained from the Electronic Communication of Surveillance in Scotland (ECOSS) with date of test and result reported. Testing for SARS-CoV-2 was carried out by real-time polymerase chain reaction (PCR) on a combined nasal and pharyngeal throat swab only in symptomatic patients. The period from 01 March 2020 to 28 February 2021 was defined as wave 1+2 whilst wave 3 commenced on 1 March 2021 and included data until 21 September 2021.

Information on hospital admissions was obtained from the Scottish Morbidity Record (SMR01) and Rapid Preliminary Inpatient Data (RAPID). Data on deaths was obtained from the National Records of Scotland (NRS). Finally, details on vaccination type and date were obtained from the Turas Vaccination Management Tool which holds all vaccination records in (TVMT) Scotland^5^. No patients were offered a third primary dose of vaccination prior to 27^th^ September 2021 and no antibody monitoring of response to vaccination was performed out with research studies.

### Definitions of outcomes

An infection was considered a ‘breakthrough infection’ if it occurred 14 days or later after receiving the second dose of vaccine. Hospitalization with SARS-CoV-2 was defined as admission within 14 days of testing positive for SARS-CoV-2 via PCR test and included those testing positive within 2 days after discharge. Death from SARS-CoV-2 was defined as death within 28 days of first testing positive for SARS-CoV-2 via PCR test.

### Statistical Methods

Data were linked and analyzed by an analyst at Public Health Scotland. Baseline characteristics of continuous variables were displayed as mean and standard deviation (SD) if normally distributed or using median and interquartile range (IQR) otherwise. Categorical variables were summarized as percentages. Crude survival was measured at 28 days since first SARS-Cov-2 test using Kaplan Meier survival curves. For each outcome (hospitalization and SARS-CoV-2 infection), the risk ratio (RR) comparing the vaccinated group with the non-vaccinated group was computed. The vaccine effectiveness (VE) was then derived from the RR using the following formula: VE=1-RR.^14^ The 95% confidence interval for VE was obtained using the nonparametric percentile bootstrap method with 10 000 repetitions. This method has been used previously for investigating vaccine effectiveness.^15, 16^ Univariable and multivariable cox proportional hazards models were fitted to examine predictors of breakthrough infections with time from 14 days post 2nd vaccine to first positive test and time from 2nd vaccine to 19/10/21 or death (which every occurred first) in those who did not test positive between 1/3/21-19/10/21.

All data were analyzed using the R statistical programming language (Version 3.6.2, Vienna, Austria).

### Ethical Statement

Formal ethical approval was waived according to Public Health Scotland Information Governance as analysis of routinely collected data. As the analysis used routinely collected and anonymized data, individual patient consent was not sought. Access and use of the data for the purpose of this work were approved following a Public Health Scotland information governance review of linking internal datasets. Only the Public Health Scotland analyst had access to the linked patient data which could only be accessed via an NHS secure network.

## Results

### Characteristics of SARS-CoV-2 positive patients

Between 1st March 2020 and 19th October 2021, there were 814 cases of SARS-CoV-2 infection in patients established on KRT, amounting to approximately 15% of the prevalent adult KRT population in Scotland. Of these, 339 (42%) were undergoing in-hospital dialysis, 29 (4%) were in home therapy and 446 (55%) were KTR. 15.6% who tested positive died within 28 days. Of those dying following a positive SARS-CoV-2 test, the majority (61%, n=78) were undergoing dialysis whilst 39% (n=49) were KTR. Table 1 shows demographics of patients by infection, dialysis modality and vaccination status. Patients dying following SARS-CoV-2 infection were older, more likely to be on dialysis, have primary renal disease of diabetic nephropathy and live in more deprived areas.

**Table 1:**
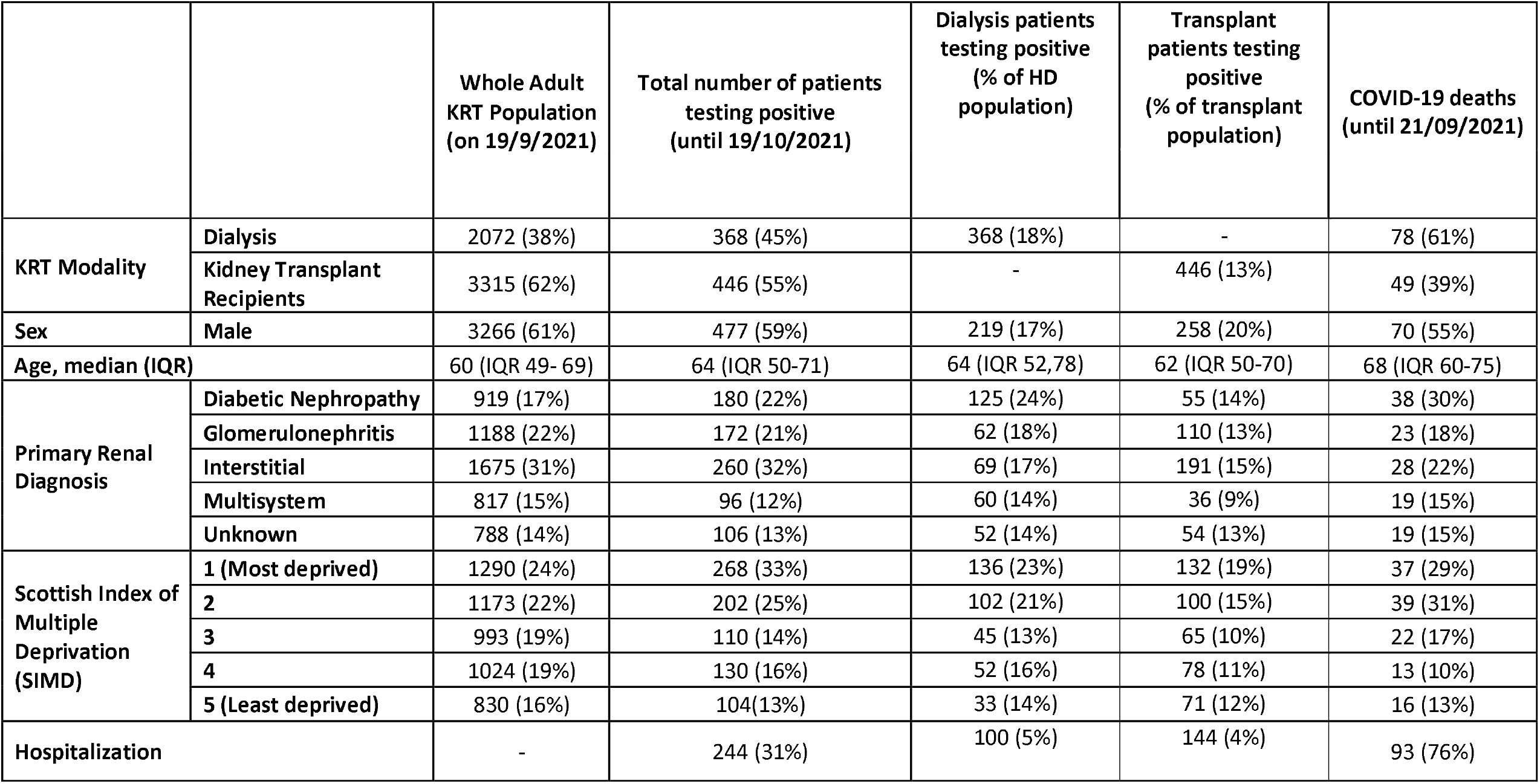
Patient demographics

Figure S1 depicts unadjusted Kaplan Meier survival curves in patients undergoing dialysis (a) and in KTR (b), stratified by wave of infection (wave 1+2 versus wave 3). In patients treated with dialysis, survival improved between waves 1+2 (survival at day 28: 0.72, 95% CI 0.68-0.78) and wave 3 (survival at day 28: 0.96, 95% CI 0.92-1.00). This was not observed in KTR, with similar survival at day 28 for wave 1+2 (0.86, 95% CI 0.82-0.92) and wave 3 (0.87, 95% CI 0.82-0.93).

### Vaccination rates

As of 19^th^ September 2021, 5281 patients (93% of the KRT population) received two doses of a SARS-CoV-2 vaccine (3522 (64%) ChAdOx1 and 1759 (29%) mRNA vaccine). 2080 patients on dialysis received at least two doses of a SARS-CoV-2 vaccine; 1283 (62%) ChAdOx1 and 797 (38%) mRNA vaccine. 3201 KTR patients received at least two vaccine doses; 2239 (70%) ChAdOx1 and 962 (30%) mRNA vaccine. Vaccination timeline in patients with chronic KRT in Scotland in presented in Figure 1a, b and c. Characteristics of vaccinated patients compared with unvaccinated patients are shown in Table 2 with unvaccinated patients more likely to be younger, male and with a transplant.

**Table 2:**
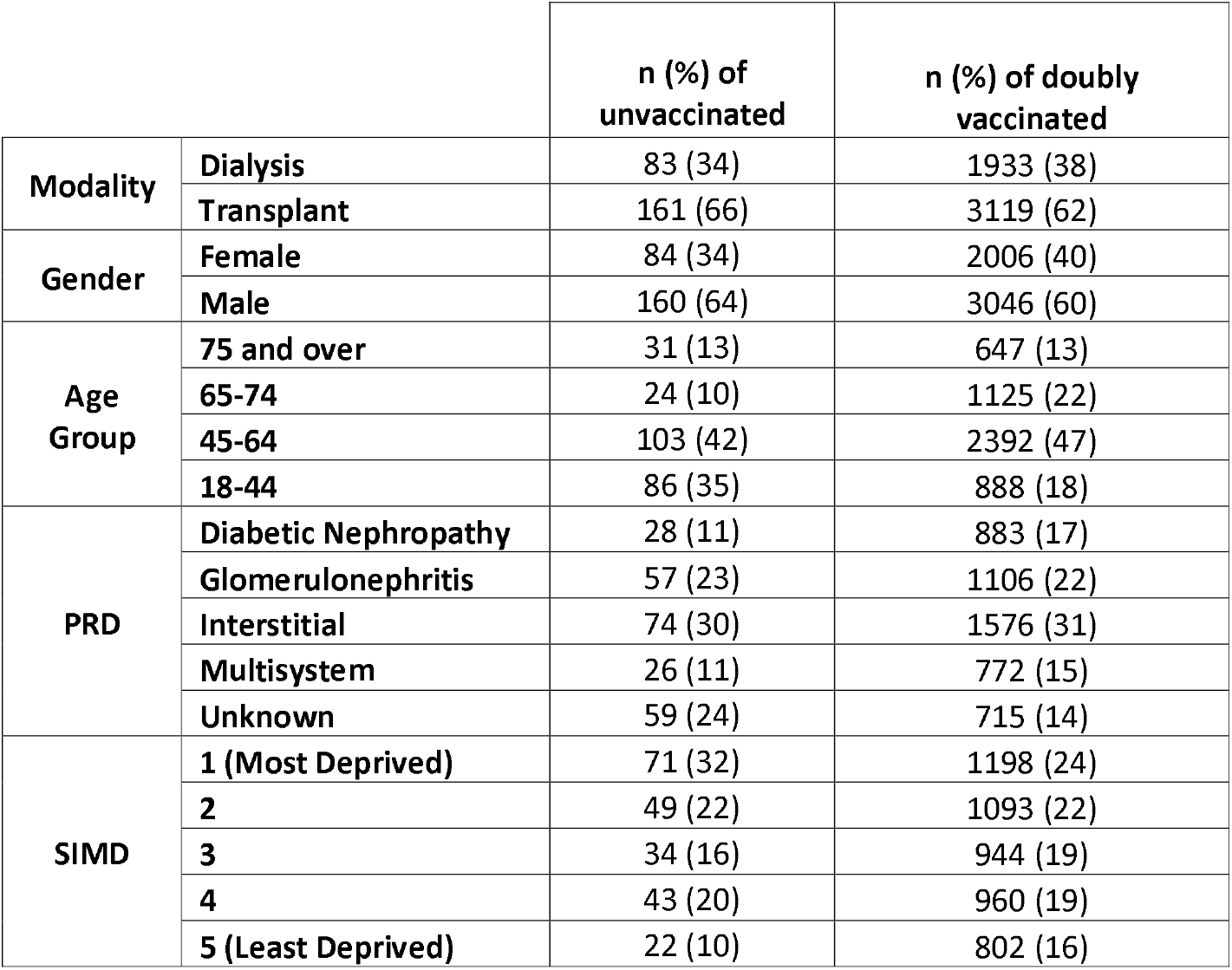
Characteristics of vaccinated compared to unvaccinated individuals

**Table 3:** Breakthrough infections (data until 19th October 2021)

|  |  | **Total number of breakthrough infections**<br>**(Positive PCR test >14 days after 2 doses)** | **Dialysis patients testing positive (with % of total prevalent HD pop)** | **Transplant patients testing positive (with % of total prevalence Tx pop)** |
| --- | --- | --- | --- | --- |
| **KRT Modality** | **Dialysis** | 98 (27%) | 98 (5%) | - |
|  | **Kidney Transplant Recipients** | 259 (73%) | - | 259 (8%) |
| **Sex** | **Male** | 210 (59%) | 58 (5%) | 152 (8%) |
| **Age, median (IQR)** |  | 57 (IQR 44-65) | 65 (IQR 54,72) | 54 (IQR 40,62) |
| **Primary Renal Diagnosis** | **Diabetic Nephropathy** | 65 (18%) | 32 (6%) | 33 (8%) |
|  | **Glomerulonephritis** | 87 (24%) | 21 (6%) | 66 (8%) |
|  | **Interstitial** | 125 (35%) | 14 (3%) | 111 (9%) |
|  | **Multisystem** | 38 (11%) | 19 (4%) | 19 (5%) |
|  | **Unknown** | 42 (12%) | 12 (3%) | 30 (7%) |
| **Scottish Index of Multiple Deprivation (SIMD)** | **1 (Most deprived)** | 108 (30%) | 32 (5%) | 76 (11%) |
|  | **2** | 84 (24%) | 28 (6%) | 56 (8%) |
|  | **3** | 54 (15%) | 13 (4%) | 41 (6%) |
|  | **4** | 57 (16%) | 16 (5%) | 41 (6%) |
|  | **5 (Least deprived)** | 54 (15%) | 9 (4%) | 45 (8%) |
| **Vaccine type** | **ChAdOx1** | 268 (75%) | 68 (6%) | 200 (9%) |
|  | **mRNA** | 89 (25%) | 30 (4%) | 59 (6%) |
| **Hospitalization** |  | 110 (34%) | 29 (1%) | 81 (2%) |
| **Death** |  | 33 (9%) | 7 (0.3%) | 26 (0.8%) |

**Figure 1a:**
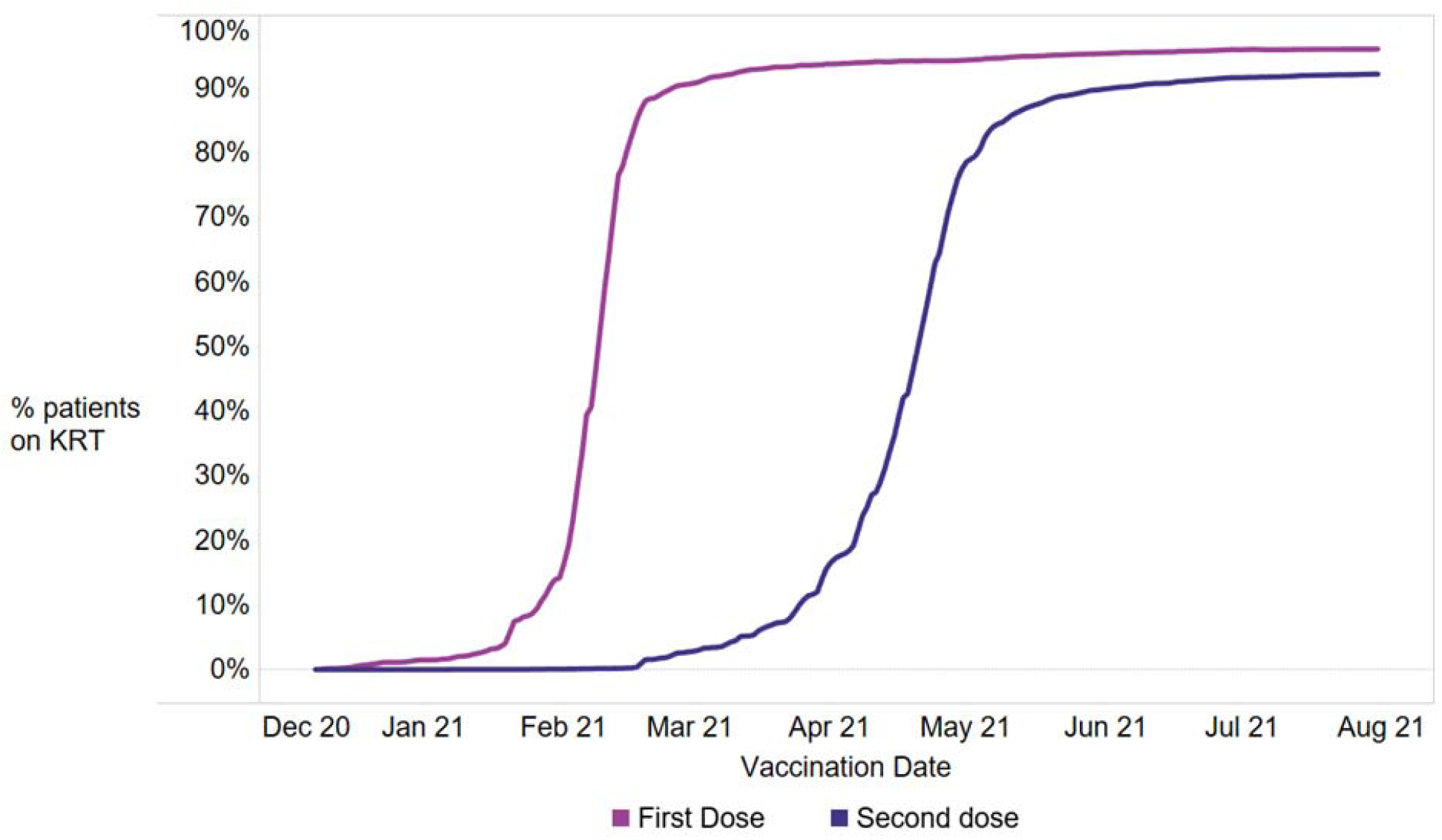
Vaccination Timeline for patients on KRT in Scotland

**Figure 1b & 1c:**
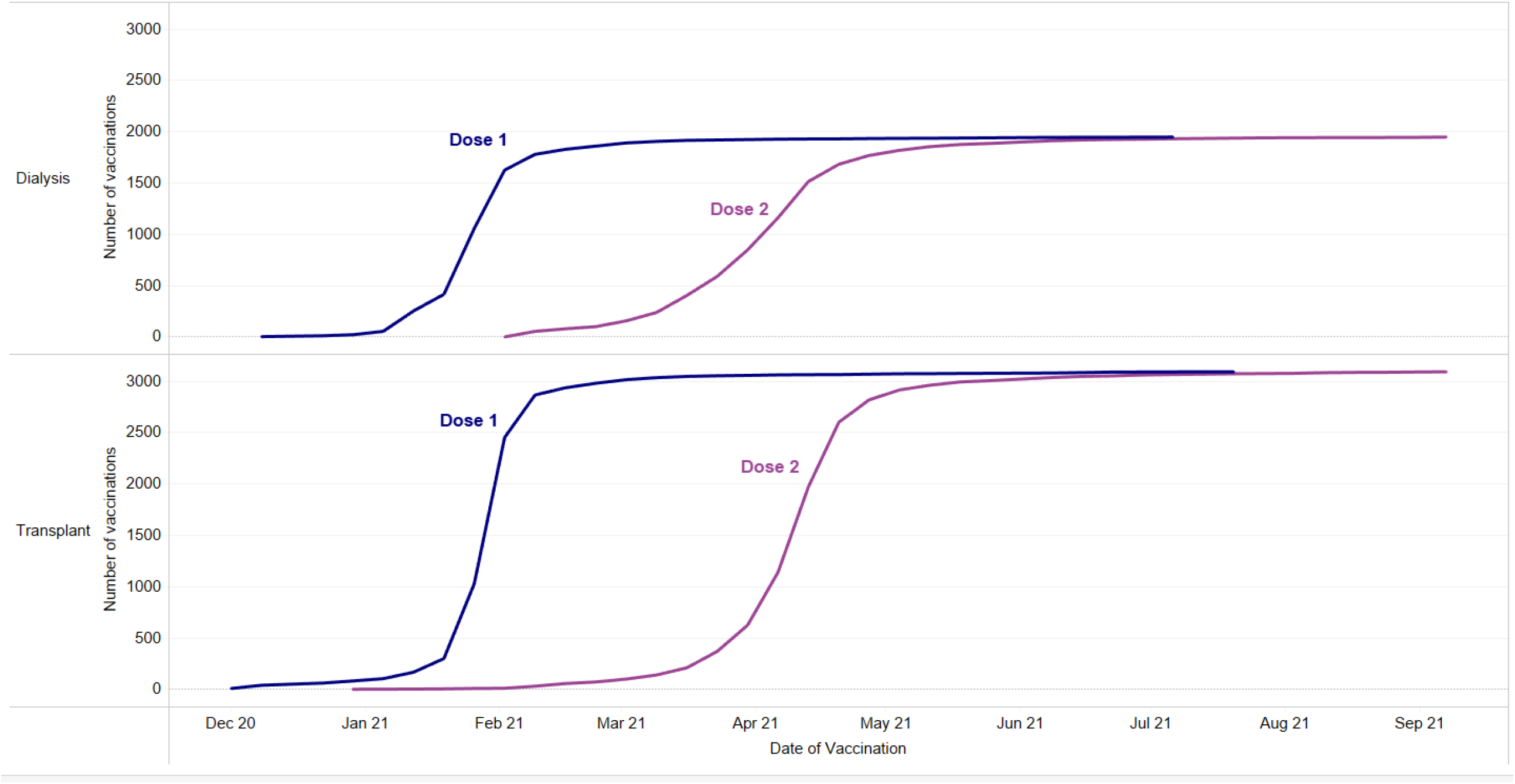
Vaccnation timline for patients receiving dialysis (b) and transplant (c)

### Vaccine effectiveness

Vaccine effectiveness against SARS-CoV-2 infection and hospitalization for any vaccine was estimated as 33% (95% CI 0-52) and 38% (95% CI 0-57) respectively (Tables S1 and S2). Vaccine effectiveness for transplant patients only was estimated at 39% (95% CI2 2-58) against infection and 40% (95% CI 0-59) against hospitalization (Tables S3 and S4). Figure S2 (a,b) displays the bootstrap distribution (histogram of bootstrap replicates) for each outcome. Until 19th October, 7.1% (n=357) patients tested positive for SARS-CoV-2 following two doses of SARS-CoV-2 vaccine (73% (n=259) ChAdOx1, 27% (n=98) mRNA vaccine). Median time from 2^nd^ dose to positive PCR was: 129 days IQR (100-140) for a mRNA vaccine; 124 days IQR (93-139) for ChAdOx1 vaccine; 131 days IQR (101-148) in dialysis patients and 123 IQR (91-137) in KTRs. Univariable and multivariable Cox proportional hazards models of predictors for breakthrough infections after 2 doses of a SARS-CoV-2 vaccine are shown in Table 4. Predictors for breakthrough infection include transplant and deprivation.

9.2% (33/357) of fully vaccinated patients who tested positive for SARS-CoV-2 died within 28 days, representing 7% (7/98) of patients undergoing dialysis and 10% (26/259) of KTR. Prior to availability of vaccination, 22.5% of patients on chronic KRT died within 28 days of a SARS-CoV-2 positive PCR test during waves 1 and 2. Overall, around 0.6% of the fully vaccinated chronic KRT population died from COVID-19, contrasting with a mortality of about 0.025% in the general Scottish population of fully vaccinated individuals (954 deaths for 3.837 million fully vaccinated people)^17^. September 2021 also saw the largest number of positive SARS-CoV-2 cases reported in the KRT population during the pandemic (Figure 2a, Figure S1a and b). This rise was greatest in the KTRs where the highest number of COVID-19 related hospital admissions was recorded since the pandemic began (Figure 2b).

**Table 4:** Univariable and multivariable cox proportional hazard model of predictors of breakthrough infection following 2 doses of SARS-CoV-2 vaccine

|  |  | HR<br>(Univariate) | HR<br>(Multivariate) |
| --- | --- | --- | --- |
| Modality | Dialysis | * | * |
|  | Transplant | 1.9 (1.5-2.5,p<0.001) | 1.8 (1.41-2.34,p<0.001) |
| Gender | Female | * | * |
|  | Male | 0.96 (0.8,1.2,p=0.7) | 0.99 (0.80-1.22,p=0.9) |
| Age | 75 and over | * | * |
|  | 65-74 | 0.96(0.60-1.54,p=0.8) | 0.81(0.51-1.31,p=0.4) |
|  | 45-64 | 1.73 (1.17-2.56,p=0.006) | 1.32 (0.88-1.98,p=0.17) |
|  | 18-44 | 2.33(1.53-3.55,p=<0.001) | 1.68(1.08-2.60,p=0.01) |
| PRD | Diabetic Nephropathy | * | * |
|  | Glomerulonephritis | 1.10 (0.80-1.52,p=0.5) | 0.98 (0.71-1.37,p=0.93) |
|  | Interstitial | 1.13 (0.83-1.53,p=0.4) | 0.98 (0.72-1.32,p=0.91) |
|  | Multisystem | 0.64 (0.42-0.96,p=0.02) | 0.68 (0.45-1.03,p=0.07) |
|  | Unknown | 0.78 (0.52-1.16,p=0.2) | 0.75 (0.50-1.13,p=0.18) |
| SIMD | 1 (Most Deprived) | * | * |
|  | 2 | 0.81 (0.60,1.07,p=0.15) | 0.81 (0.60,1.06,p=0.13) |
|  | 3 | 0.54 (0.38,0.75,p<0.001) | 0.53 (0.38-0.75,p<0.001) |
|  | 4 | 0.59 (0.43-0.82,p=0.001) | 0.56 (0.40-0.78,p<0.001) |
|  | 5 (Least Deprived) | 0.68 (0.49-0.95, p<0.02) | 0.64 (0.46,0.89,p<0.001) |

**Figure 2a:**
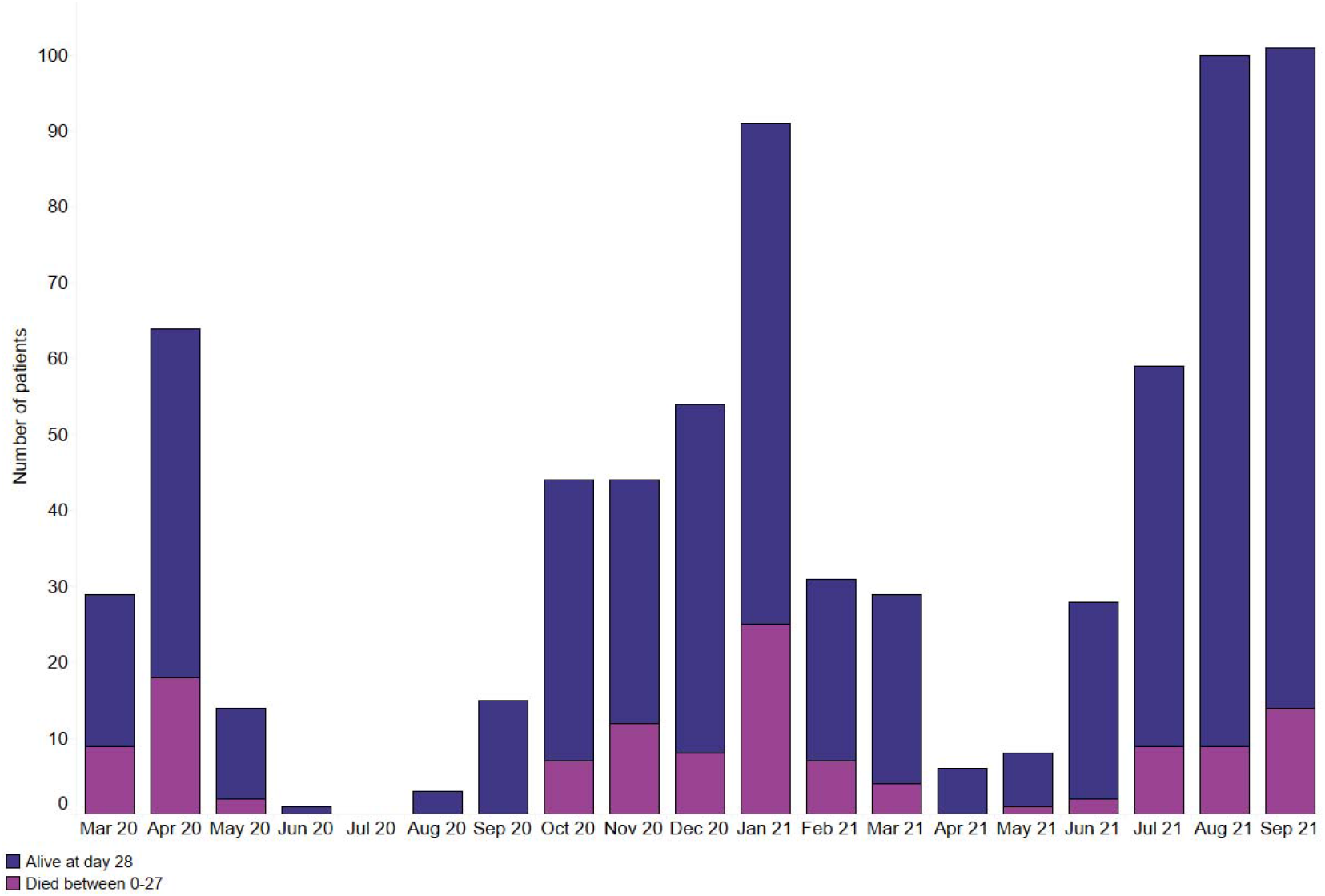
Monthly number of positive SARS-CoV-2 patients on chronic KRT in Scotland split by status at day 28 (alive in dark blue/dead in purple).

**Figure 2b:**
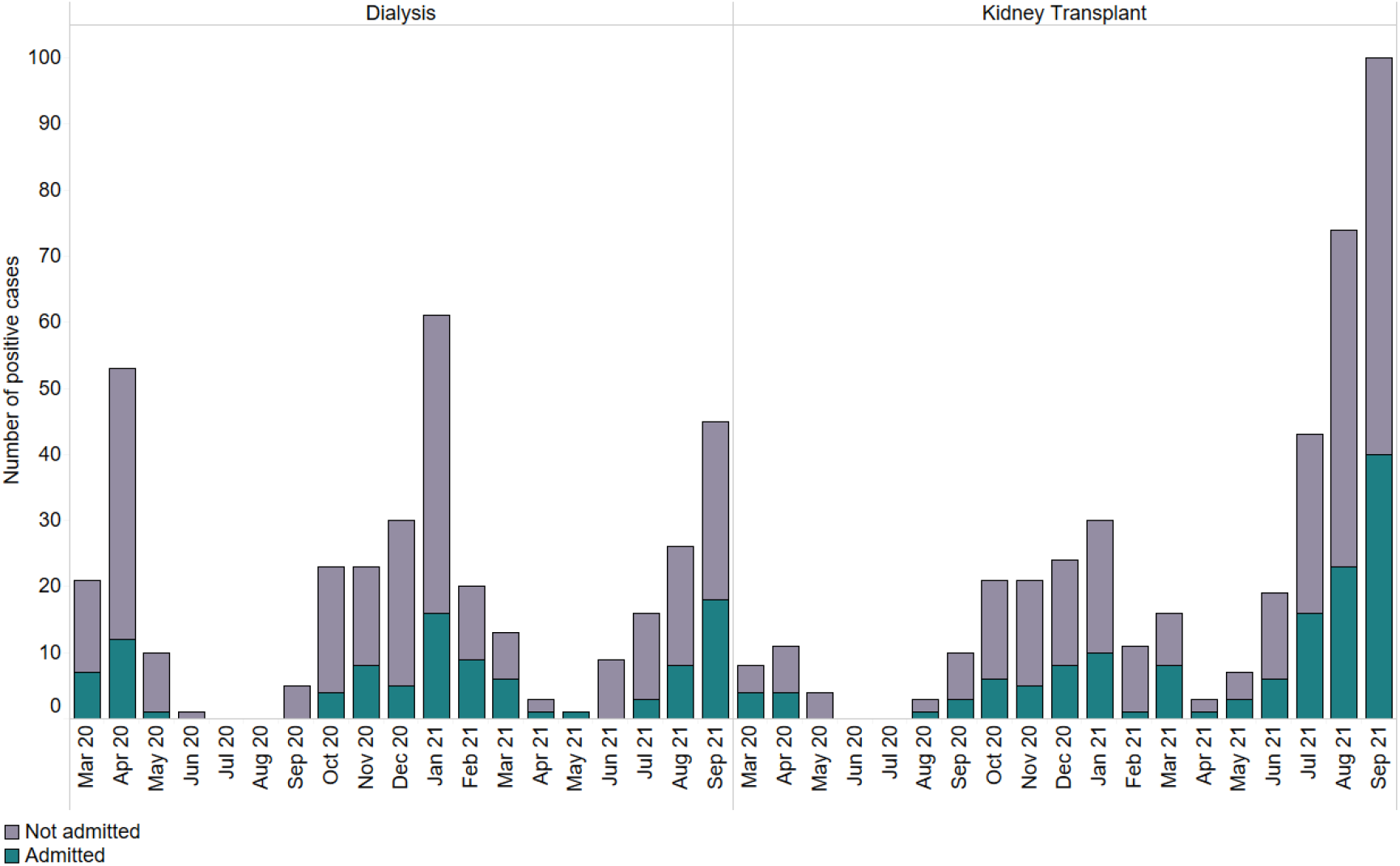

Monthly number of patients on chronic KRT testing positive for SARS-CoV-2 in Scotland split by hospitalization status (admitted in green/not admitted in grey).

## Discussion

We present data showing the impact of vaccination in patients treated with KRT in Scotland. In this group, mortality remains high following double vaccination, with 9% of patients testing positive for SARS CoV-2 dying within 28 days. Vaccine efficacy of a two-dose vaccine regimen was disappointing, with 33% reduction in risk of testing positive for SARS-CoV-2 and 38% reduction in risk of hospitalization in those testing positive, compared to the UK general population where vaccine efficacy against infection is reported as 80% (95% CI 77–83%) for BNT162b2 (Pfizer–BioNTech) and 67% (62-71%) for ChAdOx1 against the B.1.617.2 variant^18^. We found no improvement in survival between the first and latest SARS-CoV-2 infection waves in KTRs and latest data (August-September 2021) showed an alarming increase in number of SARS-CoV-2 cases and hospitalizations in this group. These data suggest that a primary vaccine course of two doses is insufficient to protect individuals on KRT. These striking adverse outcomes from COVID19 infection in KRT patients contrasts with the general population, with <0·1% of the Scottish population admitted to hospital or dying due to COVID-19 >14 days after a first COVID19 vaccine dose^19^.

We provide the first estimates of vaccine effectiveness against SARS-CoV-2 infection and hospitalization in KRT. The Scottish Renal Registry has 100% coverage of patients on chronic KRT, hence representing the epidemiology and impact of vaccination in Scotland. Our data capture a 20-month period of the pandemic including 10 months since vaccination commenced. With 93% of the KRT population ‘fully’ vaccinated in Scotland, these data allow conclusions to be drawn regarding the efficacy of a two-dose vaccination regimen.

Knowledge surrounding management of COVID-19 has increased, with interventions such as dexamethasone and tocilizumab reducing mortality in patients hospitalized with COVID-19^20, 21^. However, mortality remains high even following vaccination in KRT patients in Scotland, with 9% of patients testing positive for SARS CoV-2 dying within 28 days, much higher than in cases series reported elsewhere^22, 23^ highlighting the urgent need for adjunctive strategies to reduce risk of SARS-CoV-2 infection and its complications. It is not possible to state whether the high incidence of ‘breakthrough’ infection and adverse outcomes reflects failure to induce protective immunity, waning immunity, increasing transmissibly of the delta variant over the period of study, or wider relaxation of COVID-19 restrictions. General population data combining Brazilian and Scottish vaccination records suggests that waning immunity with ChAdOx1 significantly contributes to increased risk of hospitalization and death from COVID-19 ^24^. A large US cohort study (n=4791) demonstrated rapid waning of serological protection to SAR-CoV-2 vaccination in dialysis patients associated with risk of breakthrough infection^25^. We note much lower incidence of vaccination in the US with only 53.4% vaccinated as of 14^th^ September 2021 compared to 93% of our KRT cohort over an almost identical timescale. Strategies to address elevated risk of SARS-CoV-2 infection may include a third dose of vaccine, which induces a greater immunological response in KTR^26, 27^ and neutralizing SARS-CoV-2 monoclonal antibodies as either prophylactic or therapeutic agents.

Since completion of these analyses, the Omicron SARS-CoV-2 variant has emerged with greater propensity for evading vaccines. Booster vaccination programs have been widely instigated. Ongoing establishment of vaccine efficacy in patients requiring KRT is nevertheless required. The basic tenet of our data will remain, that patients receiving KRT are at higher risk of breakthrough infection and adverse outcomes. These patients should be prioritized for enhanced vaccination strategies and therapies such as with sotrovimab as SARS-CoV-2 neutralizing antibody with greatest proposed efficacy against Omicron ^28, 29^. Our analyses were performed prior to the widespread instigation of neutralizing antibody therapy with casirivimab plus imdevimab in patients at elevated risk of death from COVID-19^30, 31^. The role of oral antiviral therapy with molnupiravir or other agents is yet to be established, although we note that estimated GFR <30ml/min/m^2^ or requiring dialysis was an exclusion criterion in at least one clinical trial^32^.

Several limitations must be acknowledged. The numbers are too low to calculate vaccine effectiveness against death, or to separate by vaccine type. About 3% of the cohort had only received one dose of an approved vaccine at the time the data were analyzed. These were excluded from the analysis, resulting in a minor loss of statistical power. No SARS-CoV2 antibody testing was performed. Data on mortality was available until 21 September 2021 whilst SARS-CoV-2 test results were updated until 19 October 2021. Consequently, the proportion of patients dying within 28 days of their positive PCR test may be slightly underestimated. Finally, whilst it had been reported that antibody response varies according to age, degree of immunosuppression, and estimated glomerular filtration rate^8, 11, 33^, we did not conduct any adjusted analysis in this study and only report crude outcomes.

Despite promising effects of COVID-19 vaccination at a population level, for vaccinated patients requiring KRT the risk of SARS-COV-2 infection and subsequent complications remains high. Two doses of approved vaccines fail to provide sufficient protection. Further strategies to mitigate risk are required which may include non-pharmacological interventions, optimization of vaccination regimens, prophylactic therapies, and improved COVID-19 treatment strategies.

## Data Availability

The data underlying this article cannot be shared publicly as they are held by Public Health Intelligence, Scotland. Data can be requested from the electronic Data Research and Innovation Service (eDRIS) team which are part of the Public Health Scotland Public Health Intelligence, Scotland through.

## Author contributions

SB and PBM conceptualized the study; JC analyzed the data. All authors were responsible for investigation and provision of data, SB and PBM wrote the original draft; and all authors critically reviewed and edited the manuscript.

## Supplemental Table of Contents

Table S1: Vaccine Effectiveness for testing positive for COVID -19 (COVID-19 Tests Results from 01 Jan 21 – 19 October 21); Adult KRT Population as of 19 September 2021

Table S2: Vaccine Effectiveness for Hospitalization within 14 days of positive test COVID-19 Tests Results from 01 Mar 21 - 05 October 21

Table S3: Vaccine Effectiveness (Transplant patients only) for testing positive for COVID -19 (COVID-19 Tests Results from 01 Jan 21 – 19 October 21); Adult KRT Population as of 19 September 2021

Table S4: Vaccine Effectiveness (Transplant patients only) for hospitalization within 14 days of positive test COVID-19 Tests Results from 01 Mar 21 - 05 October 21

Figure S1a Monthly number of positive SARS-CoV-2 patients on dialysis in Scotland split by status at day 28 (alive in dark blue/dead in purple)

Figure S1b: Monthly number of positive SARS-CoV-2 patients with a kidney transplant in Scotland split by status at day 28 (alive in dark blue/dead in purple)

Figure S2a: Kaplan-Meier curve of 28-day survival following a positive SARS-CoV-2 PCR test in patients on dialysis in Scotland split by wave.

Figure S2b: Kaplan-Meier curve of 28-day survival following a positive SARS-CoV-2 PCR test in patients with a functioning kidney transplant in Scotland, split by wave.

Figures S3a and b: Bootstrap distribution with 95% confidence interval for relative risk of hospitalization (a) and testing positive (b) in vaccinated versus unvaccinated individuals.

