## Supplementary Material for "The impact of vaccination on incidence and outcomes of SARS-CoV-2 infection in patients with kidney failure in Scotland"

**Supplemental Material**

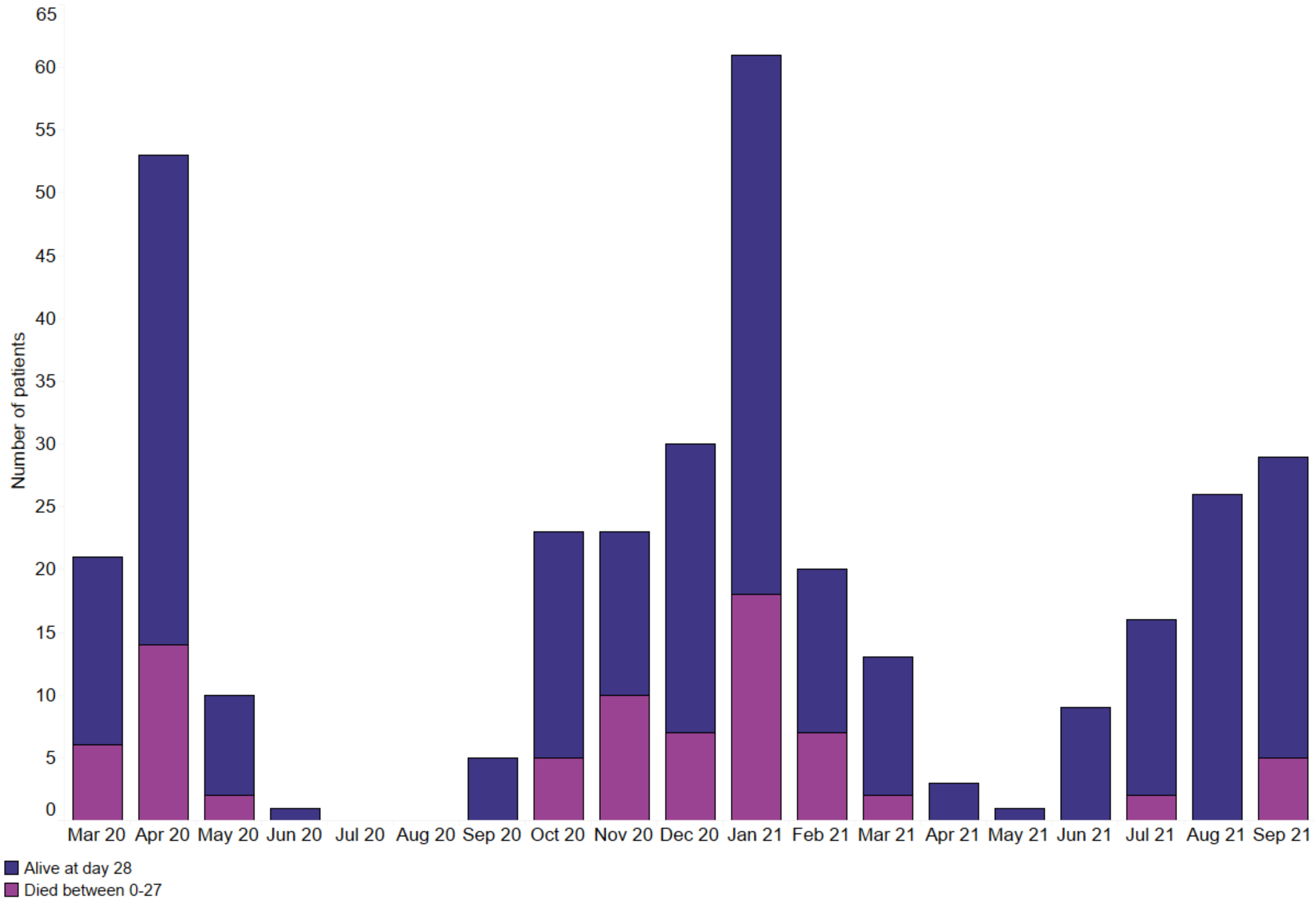

Figure S1a Monthly number of positive SARS-CoV-2 patients on dialysis in Scotland split by status at day 28 (alive in dark blue/dead in purple)

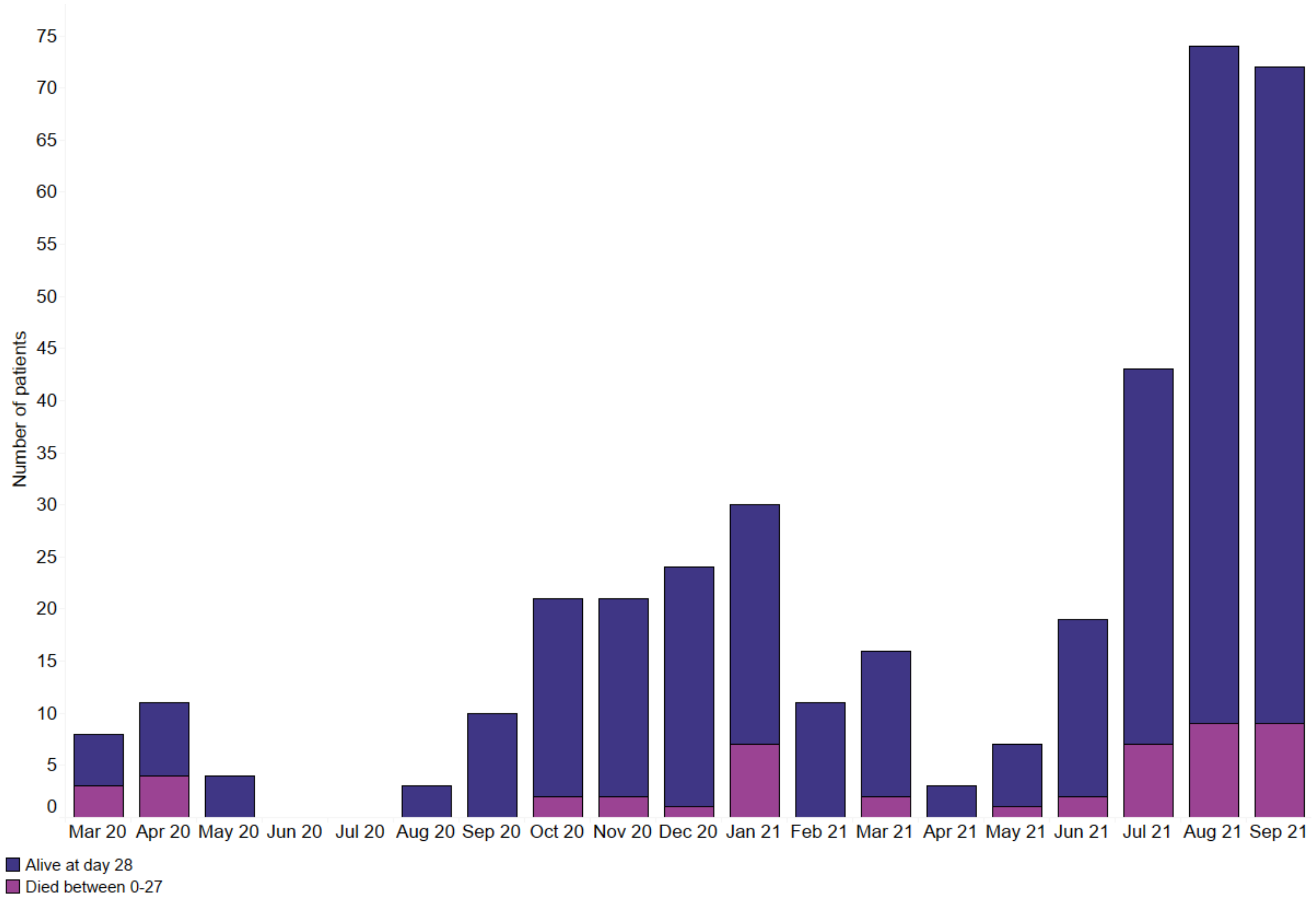

**Figure S1b:** Monthly number of positive SARS-CoV-2 patients with a kidney transplant in Scotland split by status at day 28 (alive in dark blue/dead in purple)

**Table S1: Vaccine Effectiveness for testing positive for COVID -19 (COVID-19 Tests Results from 01 Jan 21 – 19 October 21); Adult KRT Population as of 19 September 2021**

| | Positive SARS-CoV-2 | | |
|---|---|---|---|
| | **Yes** | **No** | **All Cases** |
| Vaccinated | 357 | 4654 | 5011 |
| Non-Vaccinated | 25 | 210 | 235 |
| All | 382 | 4864 | 5246 |
| | **Risk of SARS-CoV-2** | | |
| Vaccinated | 0.07 | | |
| Non-Vaccinated | 0.11 | | |
| Risk of COVID-19 in Vaccinated | 7% | | |
| Risk of COVID-19 in Non-Vaccinated | 11% | | |
| Risk Ratio | 0.67 | | |
| Vaccine Effectiveness | 33% 95% CI (0-52) | | |

**Table S2: Vaccine Effectiveness for Hospitalization within 14 days of positive test COVID-19 Tests Results from 01 Mar 21 - 05 October 21**

| | **Hospitalisation** | | |
|---|---|---|---|
| | **Yes** | **No** | **All Positive Cases** |
| Vaccinated | 110 | 216 | 326 |
| Non-Vaccinated | 12 | 10 | 22 |
| All | 122 | 226 | 348 |
| | **Risk of Hospitalisation** | | |
| Vaccinated | 0.34 | | |
| Non-Vaccinated | 0.55 | | |
| Risk of Hospitalisation in Vaccinated | 34% | | |
| Risk of Hospitalisation in Non-Vaccinated | 55% | | |
| Risk Ratio | 0.62 | | |
| Vaccine Effectiveness | 38% (95% CI 0- 57) | | |

**Table S3: Vaccine Effectiveness (Transplant patients only) for testing positive for COVID -19 (COVID-19 Tests Results from 01 Jan 21 – 19 October 21); Adult KRT Population as of 19 September 2021**

| | Positive SARS-CoV-2 | | |
|---|---|---|---|
| | **Yes** | **No** | **Total** |
| Vaccinated | 259 | 2860 | 3119 |
| Non-Vaccinated | 22 | 139 | 161 |
| All | 281 | 2999 | 3280 |
| | **Risk of Infection** | | |
| Vaccinated | 0.08 | | |
| Non-Vaccinated | 0.14 | | |
| Risk of Hospitalisation in Vaccinated | 8% | | |
| Risk of Hospitalisation in Non-Vaccinated | 14% | | |
| Risk Ratio | 0.61 | | |
| Vaccine Effectiveness | 39% | | |

**Table S4: Vaccine Effectiveness (Transplant patients only) for hospitalization within 14 days of positive test COVID-19 Tests Results from 01 Mar 21 - 05 October 21**

| | Hospitalisation | | |
|---|---|---|---|
| | Yes | No | Total |
| Vaccinated | 81 | 154 | 235 |
| Non-Vaccinated | 11 | 8 | 19 |
| All | 92 | 162 | 254 |
| | **Risk of Hospitalisation** | | |
| Vaccinated | 0.34 | | |
| Non-Vaccinated | 0.58 | | |
| Risk of Hospitalisation in Vaccinated | 34% | | |
| Risk of Hospitalisation in Non-Vaccinated | 58% | | |
| Risk Ratio | 0.60 | | |
| Vaccine Effectiveness | 40% | | |

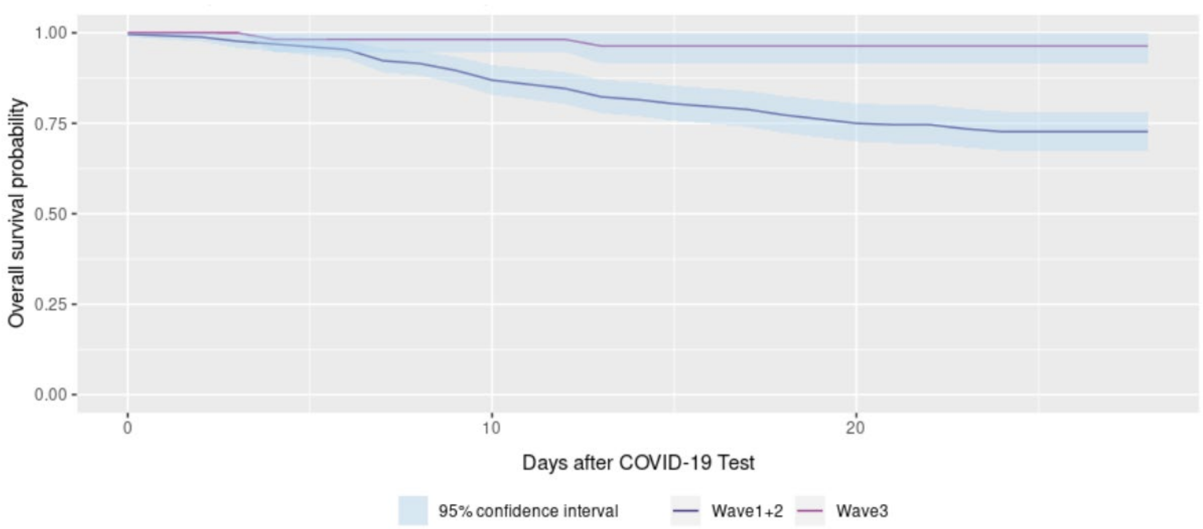

**Figure S2a**: Kaplan-Meier curve of 28-day survival following a positive SARS-CoV-2 PCR test in patients on dialysis in Scotland split by wave.

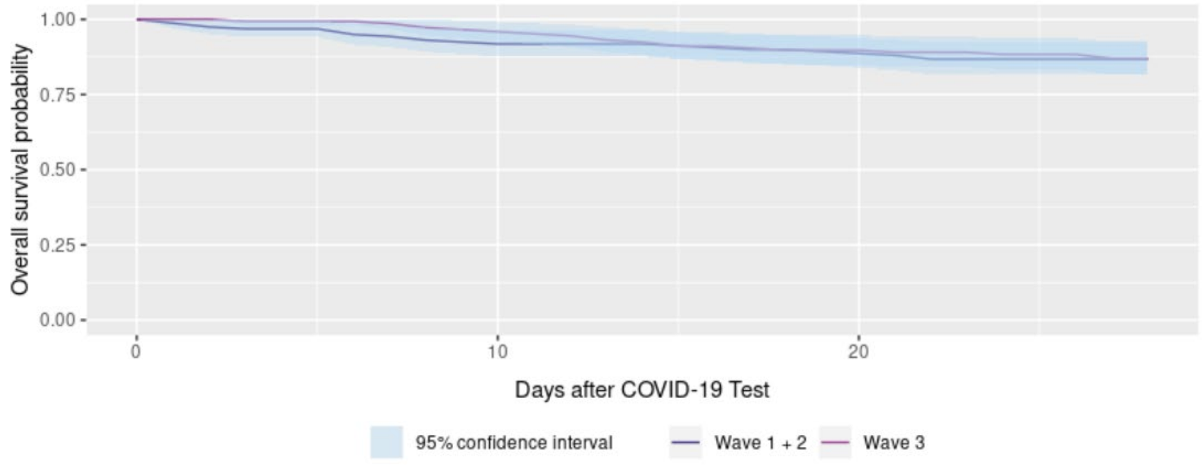

**Figure S2b**: Kaplan-Meier curve of 28-day survival following a positive SARS-CoV-2 PCR test in patients with a functioning kidney transplant in Scotland, split by wave.

a)

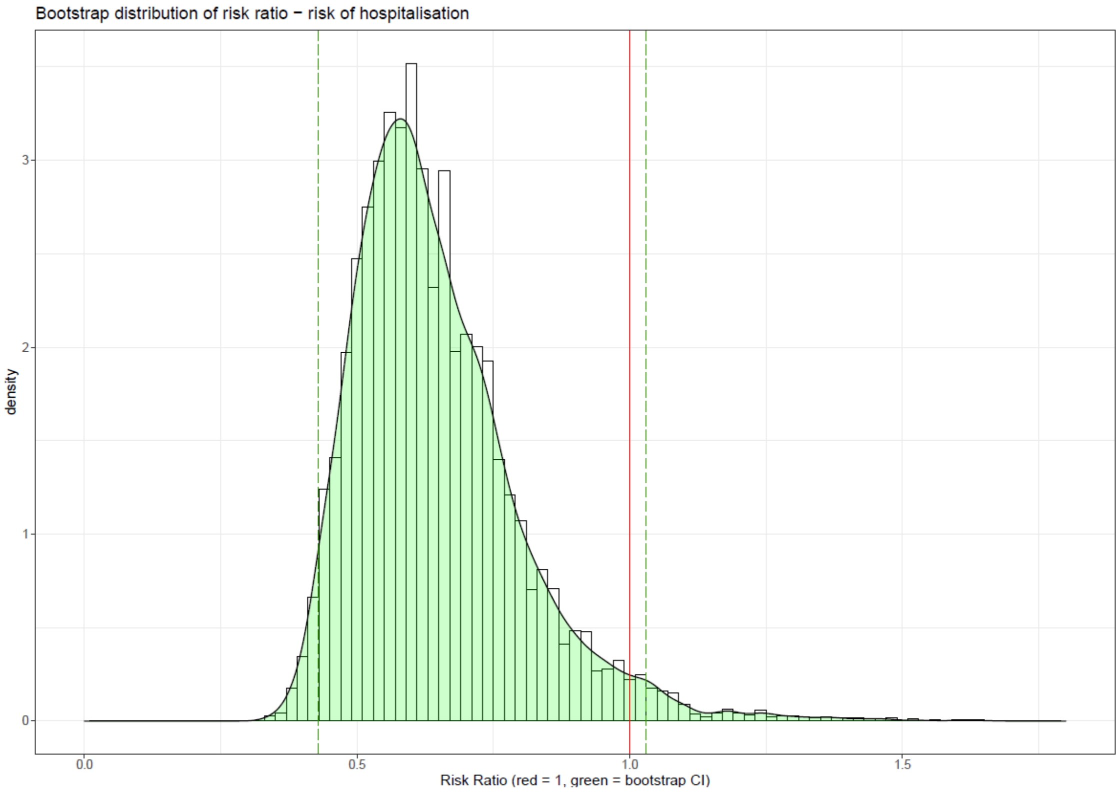

b)

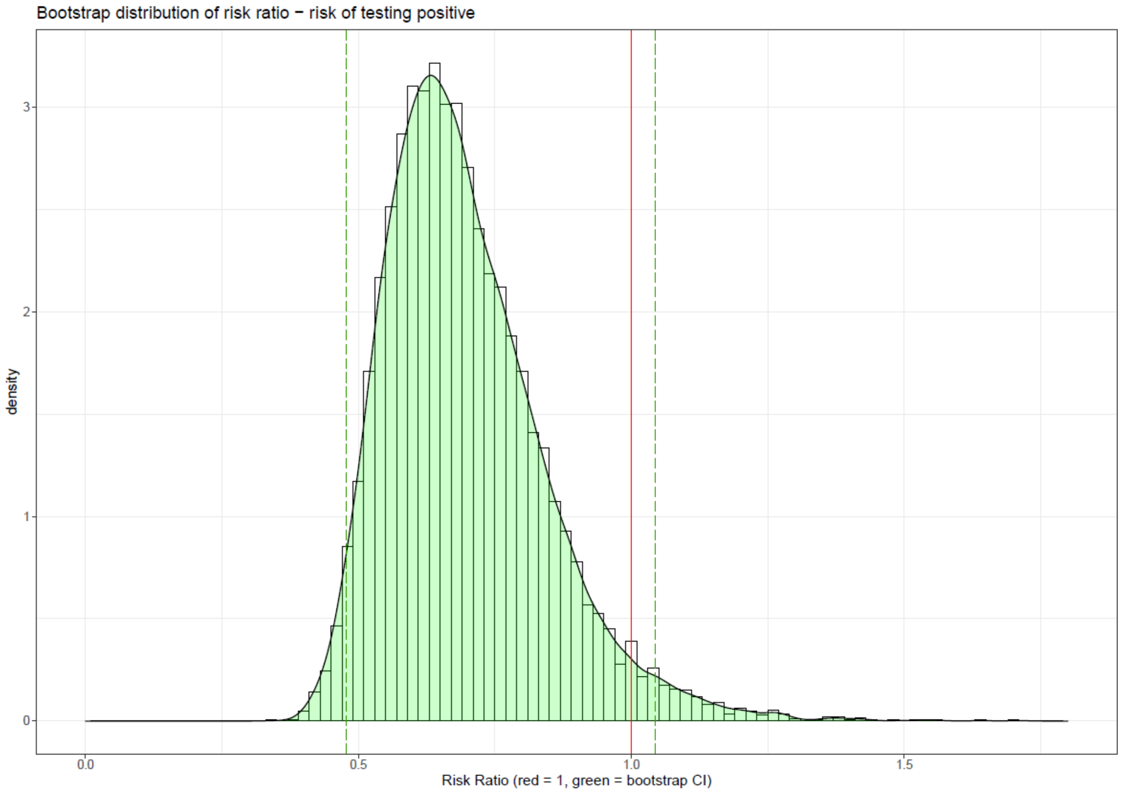

**Figures S3a and b**: Bootstrap distribution with 95% confidence interval for relative risk of hospitalization (a) and testing positive (b) in vaccinated versus unvaccinated individuals.
